# Measuring Frailty: A Comparison of the Cumulative Deficit Model of Frailty in Survey and Routine Data

**DOI:** 10.1101/2025.01.24.25321086

**Authors:** Lara Johnson, Bruce Guthrie, Atul Anand, Alan Marshall, Sohan Seth

## Abstract

**Purpose:** Frailty, a state of increased vulnerability to adverse health outcomes, impacts individuals and healthcare systems. The cumulative deficit model provides a flexible frailty measure but its application across diverse data remains underexplored. This study compares frailty indices derived from survey and routine data.

**Methods:** Frailty indices in the Clinical Practice Research Datalink (CPRD) Aurum (N = 1,625,677) and the English Longitudinal Study of Ageing (ELSA) (N = 5190) were compared for adults aged 65+ in England. Deficits were categorised as “one-to-one”, “one-to-many”, and “one-to-none”. Age-sex-standardised deficit prevalence, frailty distribution and associations with demographics were analysed using summary statistics and regression.

**Results:** Mean frailty index scores were similar (CPRD: 0.13 ± 0.10; ELSA: 0.13 ± 0.12) but differences were observed in the capture of specific deficits. The majority of deficits had a “one-to-none” or “one-to-many” mapping. Among 14 comparable deficits, visual impairment, fractures and heart failure were more common in CPRD, while falls, sleep disturbance and arthritis were more frequent in ELSA. Severe frailty and greater fitness were more prevalent in ELSA than CPRD. Sex and age influenced frailty similarly in both datasets, with frailty index scores increasing more rapidly with age in CPRD.

**Conclusion:** Differences in the number and types of deficits measured offset each other overall, supporting the cumulative deficit model’s premise that including a sufficient range of deficits does not significantly alter population-level frailty measures. This interchangeability may alleviate concerns about deficit selection, supporting more flexible approaches to population frailty assessment across both survey and routine data.

**STATEMENTS AND DECLARATIONS:** *Competing Interests and Funding:* This research was funded by the Legal & General Group (research grant to establish the independent Advanced Care Research Centre at University of Edinburgh). The funder had no role in conduct of the study, interpretation or the decision to submit for publication. The views expressed are those of the authors and not necessarily those of Legal & General. The authors have no competing interests to declare that are relevant to the content of this article.

*Ethics approval:* (1) English Longitudinal Study of Ageing: our study used anonymised data from ELSA Wave 8, which is available to researchers via the UK Data Service via an institutional log in, and we adhered to the conditions of use. The ELSA study has ethical approval and participant consent as follows: “This study involves human participants and the English Longitudinal Study of Ageing (ELSA) Wave 8 received ethical approval from the South Central – Berkshire Research Ethics Committee on 23 September 2015 (15/SC/0526) (https://www.elsa-project.ac.uk/ethical-approval). Participants gave informed consent to participate in the study before taking part. (2) The study was approved by the Clinical Practice Research Datalink Independent Scientific Advisory Committee, protocol 23_002557. We adhered to the conditions of use and did not attempt to link between datasets.

*Preprint disclosure:* This article was previously published as a preprint on medRxiv (DOI: 10.1101/2025.01.24.25321086).

**KEY SUMMARY POINTS:** *Aim:* We assess whether similar frailty prevalence is observed in survey and routine data despite differences in the number, types and prevalence of the individual deficits measured in frailty indices.

*Findings:* Frailty index calculations using established principles showed strong comparability between survey and routine data. Including a sufficient range of deficits does not significantly alter population-level frailty measures.

*Message:* The cumulative deficit model’s robustness to deficit selection supports flexible approaches to population frailty assessment across diverse data sources, such as survey and routine data.

## INTRODUCTION

Frailty is defined as a state of heightened vulnerability to adverse health outcomes compared to peers of the same age [1]. It is a significant concern for both healthcare systems and society, as it is associated with various negative outcomes, including accelerated functional decline, higher mortality, increased hospital admissions and need for long-term care [2,3,4]. Frailty’s impact extends beyond the individual, because people with frailty often require care and support which can strain personal, community and formal care resources. Frailty contributes to increased healthcare utilisation and costs, presenting challenges to health and social care systems in delivering equitable and effective care [5]. Where needs are met through informal care from family, friends and neighbours, the wellbeing and capacity of carers can be heavily impacted [6].

There are several approaches to frailty assessment, including the phenotype model (defined by the presence of clinical features) [7] and the Rockwood Clinical Frailty Scale (based on functional ability) [8]. Based on a set of physical characteristics, the phenotype model performs well but requires manual assessment and specialised equipment, making it resource-intensive and difficult to implement for entire populations. Tools like Rockwood’s Clinical Frailty Scale depend on clinicians recognising signs of frailty such as physical weakness or slowed movement. While these scores have good validity and predictive power, studies show low consistency and accuracy, and determining inter-rater reliability is challenging [9].

A difficulty of these approaches is that they rely on de novo collection of specific data, which is not feasible to do routinely or at scale. An alternative approach that can be deployed at scale using routine data is the cumulative deficit model [10]. This model quantifies frailty by counting the accumulation of “deficits” which are defined as signs, symptoms, diseases, disabilities or abnormal laboratory values [11]. A frailty index is the proportion of measured deficits present in an individual [12], facilitating population-level frailty screening using existing data. Frailty scores range from 0 to 1, where higher scores indicate greater frailty, reflecting a continuum rather than a binary state. The value of the frailty index lies in its ability to be used across different data sources, despite any differences in which deficits are measured in each source [11]. According to the cumulative deficit model of frailty, the specific nature of the deficits is less important as long as a sufficient number of them are captured [13].

The two main sources of data for frailty indices are community surveys or consented research cohorts, and linked routine data (most commonly, linked healthcare data) [11]. Surveys and research cohorts usually have the advantage of better ascertainment of psychosocial and cognitive domains, and of function, measured for example in terms of activities of daily living. Linked routine data usually has larger size, but data collection requires use of health services which may vary, and the data collected has better coverage for medically-diagnosed conditions than for social circumstances and physical and mental function. Surveys may capture more self-reported deficits by systematically asking about a wide range of potential health issues, while clinical data captures only the issues that lead someone to seek medical attention.

Our study has three aims. First, we explored whether the prevalence of individual self-reported deficits in survey data and clinically identified deficits in routine data is different. Second, we assessed whether similar frailty prevalence is observed across both data types despite differences in the individual deficits measured and the methods of data collection. Third, we examined whether demographic factors such as age and sex influence frailty scores differently in survey versus routine data.

## METHODS

### Study Design

This comparative observational study uses secondary data from two datasets to examine frailty index measures in two cohorts, with a cross-sectional analysis conducted at a single point in time.

### Data Source

This study utilised two data sources in England: Clinical Practice Research Datalink (CPRD) Aurum and the English Longitudinal Study of Ageing (ELSA). Although it is not possible to link individuals between these datasets directly, both sources cover the same geographic region and age group. However, a key difference is that ELSA excludes individuals in institutional settings, such as care homes, while GP registration and CPRD retain this population, potentially capturing higher levels of frailty.

CPRD Aurum is a longitudinal database of electronic health records from general practices in the UK [14]. The study population is people aged 65+ years and with 9+ years of permanent registration with a CPRD up-to-standard practice on 1 January 2018 (N = 1,625,677).

ELSA is a nationally representative, longitudinal survey of community-dwelling adults aged 50 years and over living in England, designed to capture the health, social, and economic circumstances of the ageing population [15]. For this analysis, we used data from 7223 Wave 8 participants, collected between June 2016 and May 2017, of whom 5190 were aged 65 years and over and had data available for at least 38 (out of 58) deficits. Participants with missing data for more than 20 deficits (N = 287) were excluded. Completeness of deficit data was high overall, with 98% of the included participants having available data on at least 57 of the 58 deficits.

### Measures

In each dataset, we developed a frailty index matching the measure used in published research: the electronic Frailty Index (eFI) in CPRD [16] and the frailty index in ELSA [17], each of which uses a distinct list of deficits appropriate to the data available.

The eFI, developed by Clegg et al, 2016, is one of the most widely used tools to measure frailty using EHRs data [18] and is automatically calculated in English GP EHRs [19]. It comprises 36 deficits [20]. Codelists containing a total of 1553 Read v2 codes [21] were used to define 35 out of the 36 eFI deficits. Read codes were mapped to medcodes, which are used for coding diagnoses in CPRD, using the EMIS Medical Dictionary. The presence of a health deficit was defined as the recording of any corresponding codes for that deficit before 1^st^ January 2018 (1 = present; 0 = absent if not recorded). The 36^th^ deficit is polypharmacy, which was defined as the prescription in the 84 days since baseline (1 January to 26 March 2010) of drugs from five or more British National Formulary (BNF) paragraphs (which define drug classes) from BNF chapters 1 to 15. Non-medications (e.g. nutritional supplements, bandages) were excluded, and missing BNF codes were imputed where necessary.

The frailty index for ELSA included 58 deficits across various domains spanning chronic diseases (e.g., hypertension), general health (e.g., vision impairment), mobility (e.g., getting up from a chair), activities of daily living (e.g., ability to get dressed), psychological wellbeing (e.g., sadness) and memory. In ELSA, deficits are systematically reassessed at each wave, so their calculation is based solely on current responses. As a result, whether a deficit was present or absent in previous waves does not influence its status in the current wave.

Frailty categories for both indices were defined based on frailty index scores, following the cut-offs proposed by Clegg et al [18]. A score below 0.12 was classified as “fit,” while scores between 0.12 and 0.24, 0.24 and 0.36, and 0.36 and above were classified as mild, moderate, and severe frailty, respectively.

### Statistical Analysis

To compare the prevalence of individual deficits between CPRD and ELSA, we computed age- and sex-standardised deficit prevalences. Individual deficits in the two frailty indices were manually mapped and categorised into three groups: “one-to-one” (directly comparable deficits), “one-to-many” (single deficits in one dataset corresponding to multiple deficits in the other), and “one-to-none” (deficits present in only one dataset). The initial mapping was performed by one author and then independently reviewed by the wider author team.

For further analysis, we selected all deficits in the “one-to-one” category. To evaluate the effect of data source on deficit prevalence, we fitted a logistic regression model for each selected deficit with age, sex (0 = male, 1 = female), and data source (0 = ELSA, 1 = CPRD) as independent variables. Based on this model, we calculated age-sex-adjusted odds ratios for the data source. The model was specified as:

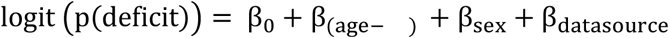

where β_o_ is the intercept and we have used β_x_ to denote β_x_ × *x* for simplicity. To assess population differences in the prevalence of frailty, we inspected the distribution of frailty scores in CPRD and ELSA. Frailty scores were calculated and stratified by age group, and we initially compared the distributions using histograms and summary statistics. Because of non-normality of frailty scores, to statistically evaluate any differences we used the Kolmogorov-Smirnov (K-S) test to compare the overall distributions, and calculated the median and interquartile ranges.

To explore associations between demographic factors and frailty scores we used summary statistics and employed a linear regression model with age, sex and data source as independent variables. The regression equation used was:

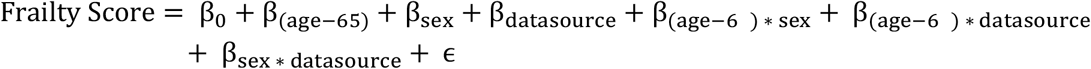

Analysis was performed using Python (version 3.11.3) and the *statsmodels* and *scikit-learn* libraries.

## RESULTS

The study population comprised 1,625,677 individuals from the CPRD 2018 Cohort and 5190 from ELSA Wave 8, all aged 65 years and older (Table 1). The proportion of women was 53.6% in CPRD and 56.3% in ELSA. The mean age was 75.5 years in CPRD (SD = 7.7) and 75.0 years (SD = 7.1) in ELSA. 48.3% of individuals in CPRD had mild, moderate or severe frailty (defined as an eFI score >= 0.12), compared to 42.8% in ELSA.

**Table 1.** Study population characteristics of individuals over age 65 years.

| Variable | CPRD 2018 Cohort |  | ELSA Wave 8 |  |
| --- | --- | --- | --- | --- |
|  | N | % | N | % |
| **Subjects** | 1,625,677 |  | 5190 |  |
| **Age in Years (Mean ± SD)** | 75.5 ± 7.7 | | 75.0 ± 7.1 | |
| 65-69 | 420,506 | 25.9% | 1407 | 27.1% |
| 70-74 | 425,675 | 26.2% | 1397 | 26.9% |
| 75-79 | 305,879 | 18.8% | 980 | 18.9% |
| 80-84 | 237,565 | 14.6% | 853 | 16.4% |
| 85-89 | 148,817 | 9.2% | 368 | 7.1% |
| 90+ | 87,235 | 5.4% | 185 | 3.6% |
| **Sex** |  |  |  |  |
| Men | 754,427 | 46.4% | 2270 | 43.7% |
| Women | 871,250 | 53.6% | 2920 | 56.3% |
| **Frailty index score (Mean ± SD)** | 0.13 ± 0.10 | | 0.13 ± 0.12 | |
| Fit (< 0.12) | 840,443 | 51.7% | 2971 | 57.2% |
| Mildly frail (≥ 0.12 and < 0.24) | 531,961 | 32.7% | 1300 | 25.0% |
| Moderately frail (≥ 0.24 and < 0.36) | 197,668 | 12.2% | 580 | 11.2% |
| Severely frail (≥ 0.36) | 55,605 | 3.4% | 339 | 6.5% |

### Individual Deficit Prevalence and Comparison

To compare the individual deficits in CPRD and ELSA, we inspected standardised deficit prevalence by sex and age group (Supplementary Table 1). Overall, the standardised deficit prevalence was similar in CPRD (mean = 13.7%; SD = 12.9%) and ELSA (mean = 13.1%; SD = 11.8%). Prevalence ranged from 0.7% (activity limitation) to 51.4% (hypertension) in CPRD and 0.6% (Alzheimer’s) to 48.9% (arthritis) in ELSA.

The deficits included in the two indices can be broadly categorised in three groups (Figure 1):

**Figure 1.**
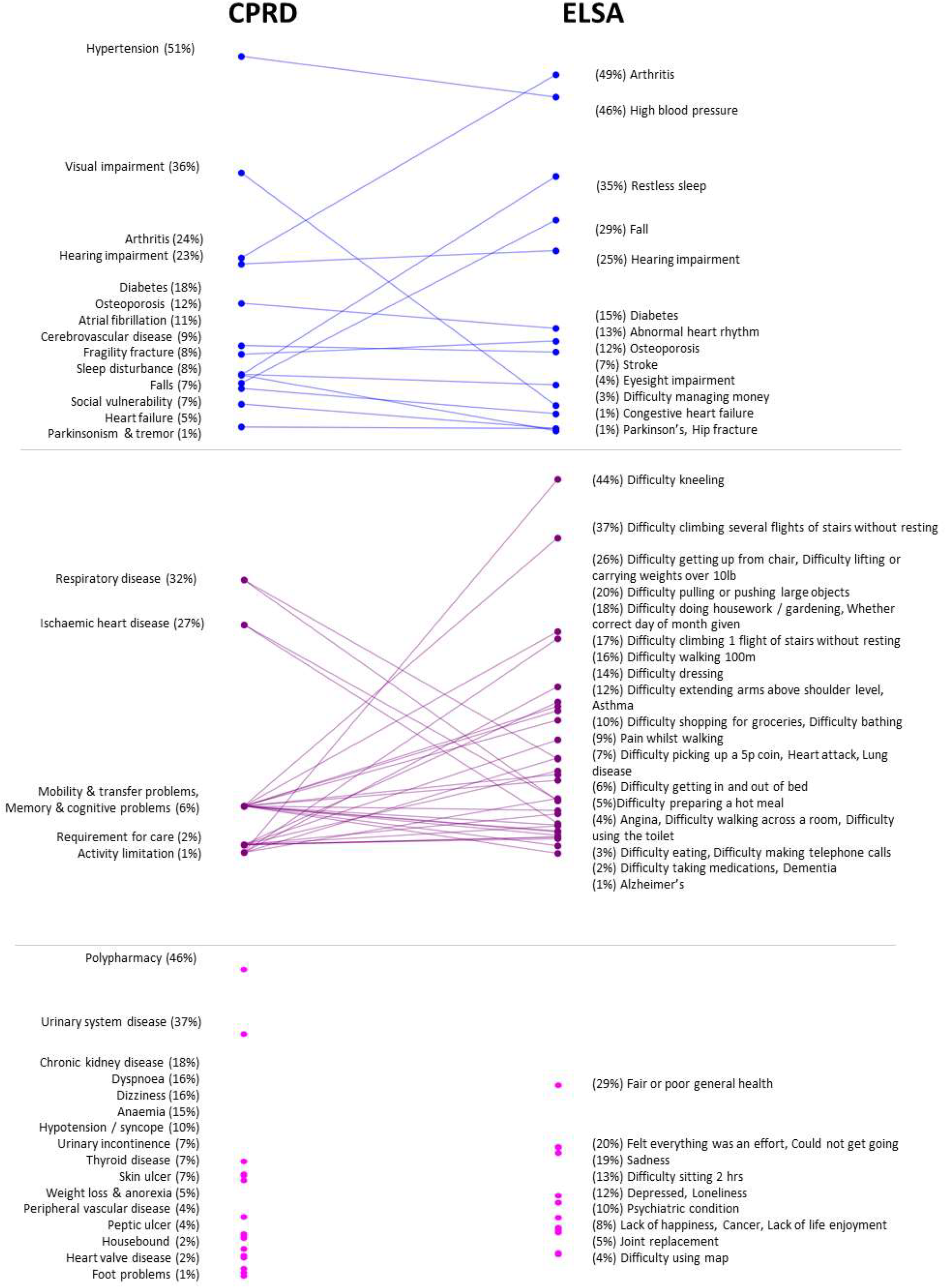
Deficit Prevalence Comparison between CPRD and ELSA. Each point represents a health deficit, with prevalence in CPRD (left) and ELSA (right). Blues line indicate a one-to-one mapping, purple lines one-to-many and pink points a one-to-none. For full details on each health deficit and its mapping across datasets, see Supplemental Table 1. Figure footnote: Items related to “Memory & cognitive problems” in CPRD overlap with “Mobility & transfer problems” and do not appear as separate points.

1. **Deficits with similar definitions (one-to-one)**: 14 deficits were classified as directly comparable based on conceptual and clinical similarity. For example, “Diabetes or high blood sugar” in ELSA maps to “diabetes” in CPRD. This category comprised morbidities, falls and fractures. While exact equivalence between self-reported and clinically recorded deficits is rarely achievable, the deficits in this category were considered to reflect the same underlying health construct. Nonetheless, even among these, differences in definition remain - for instance, sleep disturbance in CPRD indicates a GP consultation for sleep disturbance, whereas in ELSA it can refer to self-reported restless sleep during the previous week.
2. **Deficits corresponding to multiple deficits (one to many)**: In some cases, a single deficit in one dataset corresponded to multiple deficits in the other. Six broader deficits in CPRD mapped to 31 more detailed deficits in ELSA. For example, the “mobility and transfer problems” deficit in CPRD was related to several specific mobility deficits in ELSA, such as “difficulty walking 100m” and “difficulty climbing one flight of stairs without resting”. Similarly, the “requirement for care” deficit in CPRD was related to several activities of daily living deficits in ELSA, including “difficulty dressing” and “difficulty eating”. This category predominantly included mobility impairments, physical limitations and memory problems, which were recorded in more granular detail in ELSA than in CPRD. However, some morbidities also fell into this category. For example, “lung disease” and “asthma” in ELSA mapped to the broader “respiratory disease” deficit in CPRD.
3. **Unique deficits (one to none)**: Some deficits were unique to each dataset. There were 13 deficits found only in ELSA, while 16 deficits were unique to CPRD. ELSA included several mental health deficits not captured in CPRD, such as “sadness”. Conversely, CPRD included the polypharmacy deficit, which was absent in ELSA.

For the 14 deficits that had similar definitions in both datasets, we inspected the age-sex adjusted odds ratios (ORs) (Figure 2 and Supplementary Table 2). Eight deficits were more likely in CPRD than ELSA, including visual impairment (13.50), fragility fracture (10.38), heart failure (3.77), social vulnerability (2.07), diabetes (1.26), hypertension (1.22), cerebrovascular disease (1.15) and osteoporosis (1.10). Five deficits were less likely in CPRD than ELSA, including falls (0.16), sleep disturbance (0.17), arthritis (0.32), atrial fibrillation (0.78) and hearing impairment (0.86). Only “Parkinsonism & tremor” did not have a statistically significant difference at a threshold of p < 0.05.

**Figure 2.**
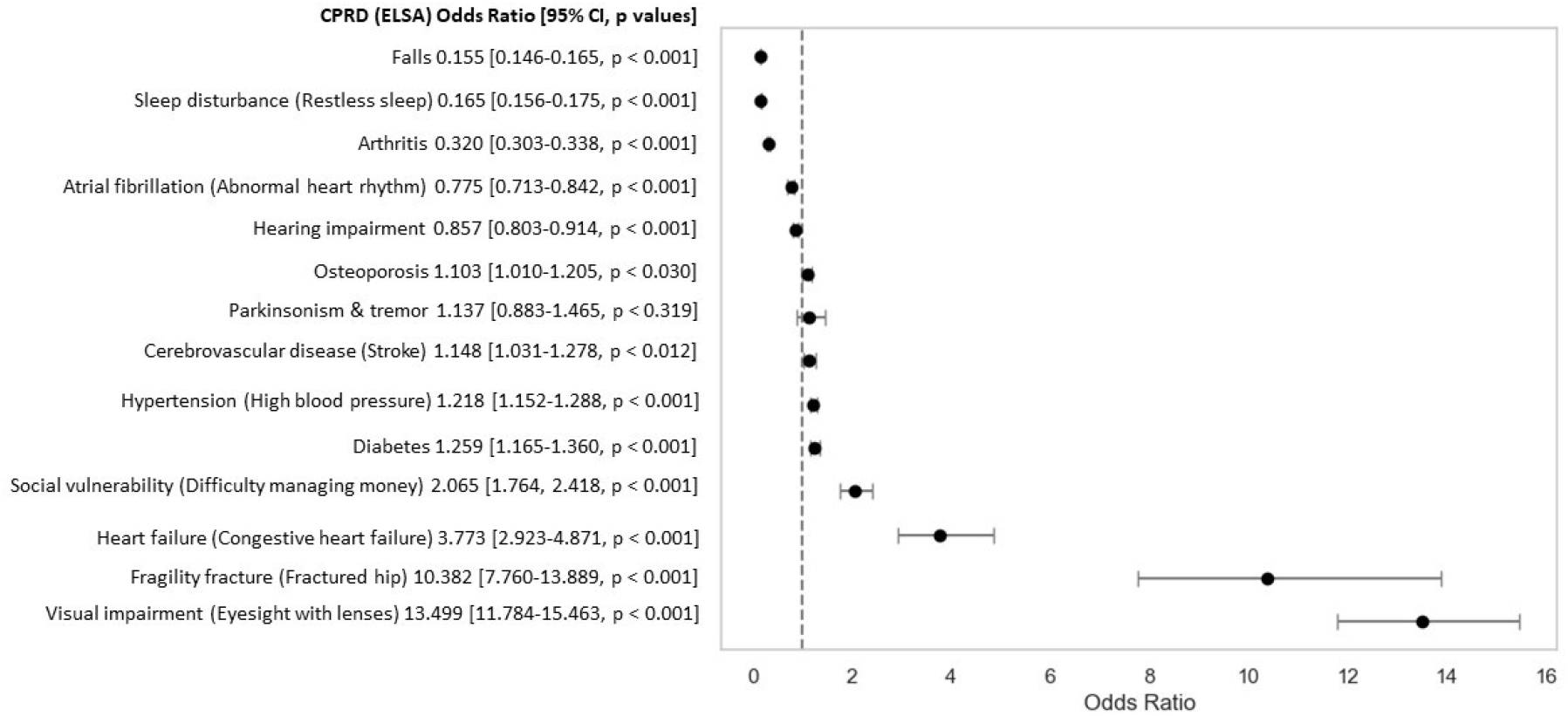
Forest Plot of Age-Sex Adjusted Odds Ratios for Deficits with Similar Definitions in ELSA and CPRD. Deficits plotted to the left of the vertical line (OR = 1) are more prevalent in ELSA, while those to the right are more prevalent in CPRD.

### Population-level Frailty Prevalence and Distribution

To assess population differences in the prevalence of frailty, we visualised the frailty score distributions per data source using a histogram (Figure 3) and a plot of frailty scores by age group (Figure 4, Supplementary Figures 1 and 2), along with summary statistics. ELSA values were more skewed in all age groups than CPRD. The Kolmogorov-Smirnov (K-S) test produced a statistic of 0.176 (p < 0.001), indicating a statistically significant difference in the overall distributions. We expect this since ELSA contains more individuals at either spectrum of the frailty values compared to CPRD. While mean frailty index scores were similar between sources (ELSA: 0.13 ± 0.12; CPRD: 0.13 ± 0.10), the median score was higher in CPRD at 0.138 (IQR 0.056-0.194) than in ELSA at 0.086 (IQR 0.052-0.190), p < 0.001. The maximum score was 0.74 in ELSA and 0.75 in CPRD.

**Figure 3.**
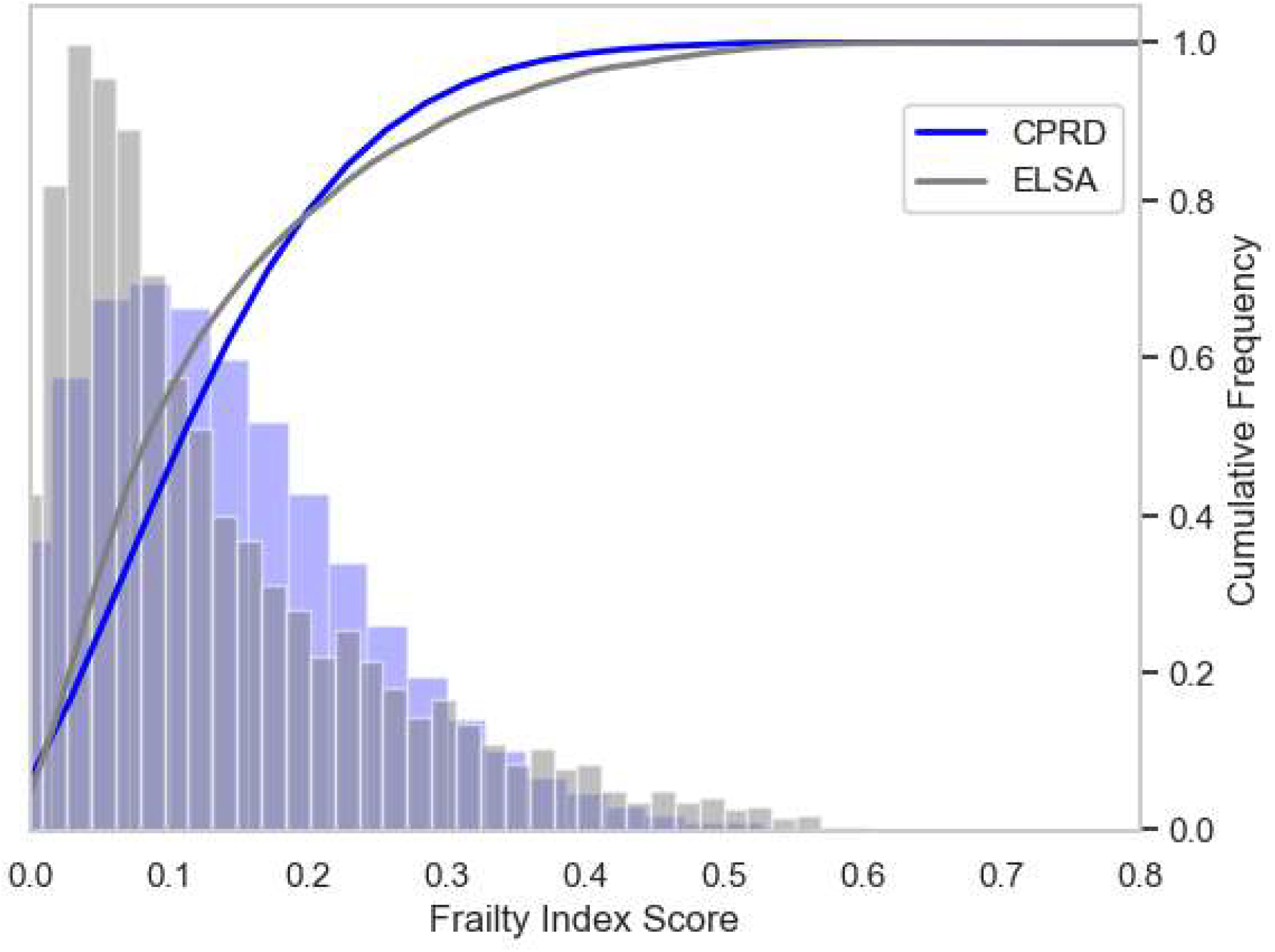
Histogram and Empirical Cumulative Distribution Function (ECDF) Plots of Frailty Scores, with 36 bins for CPRD and 58 bins for ELSA. The histogram was normalised to account for differences in bin counts across data sources.

**Figure 4.**
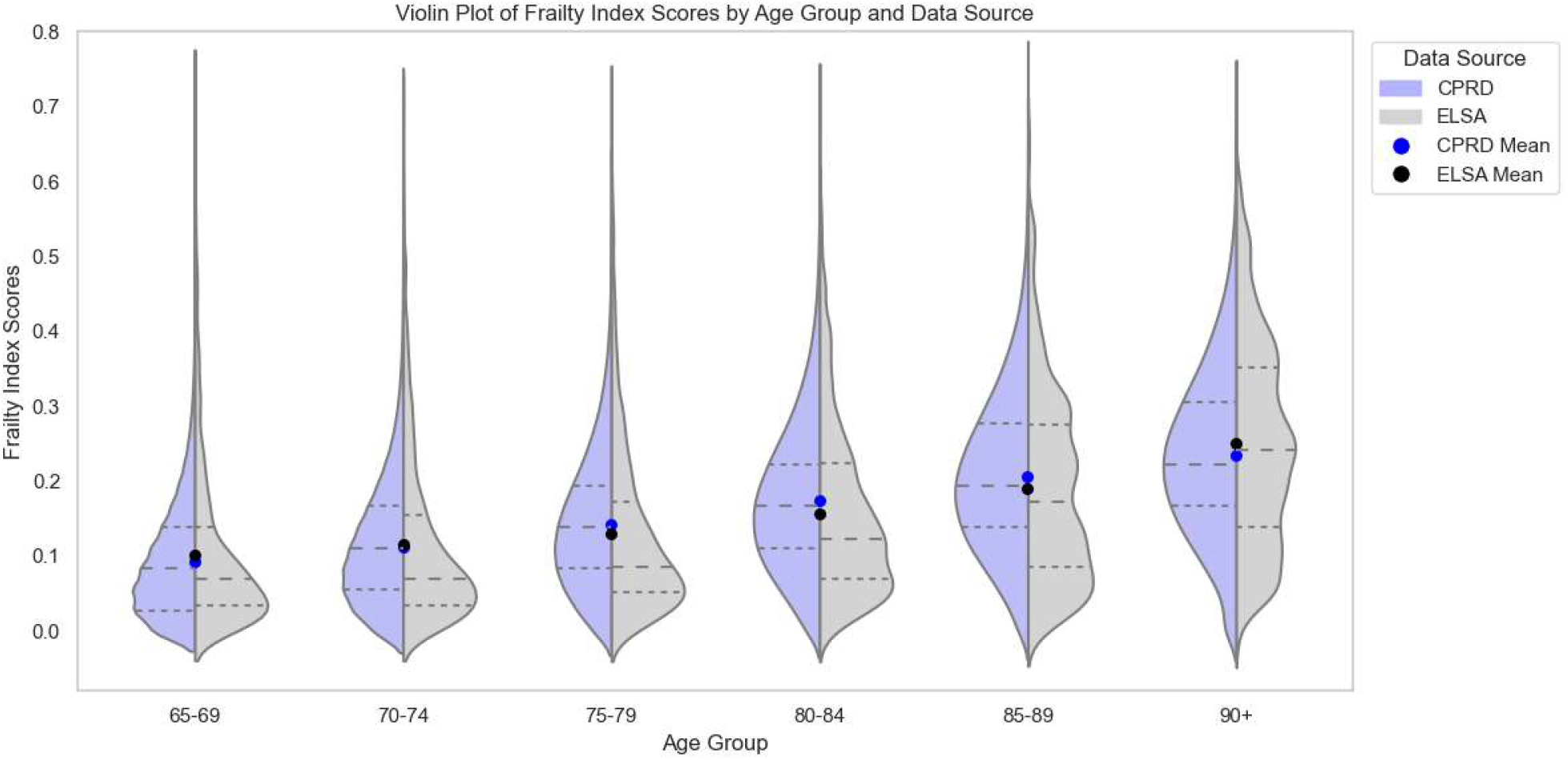
Frailty Scores by Age Group and Data Source. Dashed lines represent the first quartile (Q1), median (Q2), and third quartile (Q3) of frailty scores within each age group.

### Demographic factors

The linear regression model predicting frailty index scores (Supplementary Table 3 and Supplementary Figure 3) showed that both age (β_()_=0.0046, p<0.001) and female sex (β_s x_=0.0241, p<0.001) were positively associated with frailty scores. The CPRD data source was not significant on its own (β_d t sourc_ =-0.0029, p < 0.245). Interaction terms further reveal that the relationship between age and frailty is moderated by both sex and data source; specifically, frailty increases more steeply with age for women in ELSA (β_(age-65)*sex_ = 0.0002, p < 0.001) and within CPRD for men (β_(age) ∗ datasourc_ = 0.0009, p < 0.001). Additionally, the difference in average frailty between men and women aged 65 are less pronounced in CPRD than in ELSA (β_sex ∗ datasourc_ = −0.0163, p < 0.001). The model explained approximately 19.8% of the variance in frailty index scores (R^2^ = 0.198, p < 0.001).

## DISCUSSION

We have compared the application of established frailty indices to ELSA survey and CPRD routine GP records, noting that while mean index scores were similar across these cohorts, important differences were observed in the capture of specific deficits. The CPRD cohort included more adults in the oldest age group compared to ELSA, but the frailty index in ELSA identified more people as severely frail. Among 14 directly comparable deficits, visual impairment, fractures and heart failure were more commonly seen in routine GP data, while falls, sleep disturbance and arthritis were more often reported by survey participants. The majority of deficits either did not map between CPRD and ELSA or had a one-to-many mapping. Overall frailty index scores appeared to increase more steeply with age in the CPRD dataset, but with a smaller difference between men and women than observed in ELSA.

A key advantage of the frailty index is its flexibility. Established guidelines for constructing a frailty index are grounded in principles designed to capture age-related declines across various body systems, incorporating robustness checks on their distributions [11] but do not predetermine specific deficits. This flexibility has led to a wide range of indices with varying items and item counts based on data availability [22,23]. While systematic reviews have examined population-level frailty prevalence [24] and frailty-outcome relationships [25], detailed comparisons of individual deficits across indices are still lacking, leaving a gap in understanding how specific deficit choices impact frailty measurement.

Differences in individual deficit prevalence between routine and survey data may arise from several factors. Routine data, capturing individuals who interact with healthcare services, may have a disease-predominant coding bias, reflecting the role of the GP and coding practices, as opposed to more activity-based measures. Differences in treatment-seeking behaviour also play a role; for example, arthritis was reported twice as often in ELSA, likely reflecting self-reported joint pain as “arthritis” without a formal diagnosis. Medical data may underreport disabilities [26], sleep issues, or falls. Varying definitions also contribute; for instance, it is clear from the prevalence of visual impairment in CPRD that it lacks specificity for significant sight loss and is likely to include large numbers with minimal issues such as needing glasses. ELSA defines eyesight impairment more narrowly as difficulty seeing even with glasses. Within routine data, prevalence differences can also occur; for example, myocardial infarction rates in CPRD were underestimated by 25-50% using one source compared to using GP and Hospital Episode Statistics (HES) data [27], showing a need for both to truly capture morbidity in routine data [28].

In CPRD, the mean frailty index score is 0.13 (SD = 0.10), which closely aligns with the 0.14 (SD = 0.09) observed in the ResearchOne primary care database in England, where the eFI was originally developed [18]. The distribution of frailty categories is also similar: in CPRD, 51.7% are classified as fit, 32.7% as mildly frail, 12.2% as moderately frail, and 3.4% as severely frail, closely matching the proportions reported in Clegg et al. (50% fit, 35% mild, 12% moderate, 3% severe) [18]. These findings suggest that the overall prevalence and distribution of frailty in CPRD are broadly consistent with other UK-based studies.

The eFI in CPRD and the frailty index in ELSA produce comparable population-level frailty estimates, with similar mean scores and distributions resembling a gamma distribution and a natural upper limit of < 0.7 based on visual inspection of the histograms. However, the distributions are statistically distinct, with ELSA showing a greater proportion of individuals at both the lower and upper ends of the frailty scale. The higher frequency of very low frailty scores in ELSA may stem from its larger set of 58 deficits (compared to 36 in CPRD), meaning a higher number of deficits is required to reach the same index score. Conversely, ELSA’s higher rate of severe frailty scores may result from a wider range of assessed deficits, including social and environmental factors absent in clinical records. This pattern persists even though ELSA excludes care home residents, who typically exhibit higher frailty levels, and includes fewer people in the oldest age groups. These differences in score distribution have important implications for the application of frailty categories. A threshold-based classification may yield different prevalence estimates depending on the data source, even when the underlying mean frailty index score is similar. For instance, ELSA may classify more individuals as severely frail due to the inclusion of self-reported deficits, whereas CPRD may under-identify these individuals. This highlights the need to consider context and deficit composition when interpreting frailty categories across datasets, particularly in research or policy settings where such classifications inform risk stratification, resource allocation, or clinical decision-making.

This study benefits from the use of established frailty indices applied to representative samples of the English population [27,29]. Comparing the eFI in CPRD with the frailty index in ELSA allows for meaningful insights into the variability of frailty measures across two distinct data types at the population level. However, a key limitation is the lack of linked data, that would enable calculating both the frailty index and eFI for the same individuals, preventing direct, individual-level comparisons between the two measures. Additionally, while ELSA has agreements with NHS Digital to link mortality data and Hospital Episodes Statistics, this data is not yet accessible to external researchers, limiting the potential for evaluating frailty measures against health outcomes [30]. Finally, the lack of socioeconomic or multiple deprivation index data in CPRD GP data restricts the ability to contextualise frailty within broader social determinants.

Our findings suggest that, despite variations in the number and types of deficits measured, these differences offset each other overall, supporting the cumulative deficit model’s premise that including a sufficient range of deficits does not substantially alter the frailty measure [13]. This interchangeability may alleviate concerns about deficit selection, supporting more flexible, holistic approaches to frailty assessment across varied health domains. Such flexibility is valuable, as focusing solely on medical conditions may overlook critical cognitive, social, and psychological aspects [31, 32].

In conclusion, while comparison of individual items is problematic across ELSA and CPRD, where frailty index statistics are calculated using established principles there is a high level of comparability at a population level. This finding is in line with the theory and principles that underpin the frailty index construction and so researchers can have some confidence in comparing results across datasets. This might stimulate research on frailty that draws on both social survey and routine data (and encourages linkages between them) given complementarities in strengths and weaknesses.

## Data Availability

All data produced in the present work are contained in the manuscript.

## Acknowledgements

We thank Professor Andrew Clegg, who generously shared the eFI codelists.

## CONFLICT OF INTEREST STATEMENT

On behalf of all authors, the corresponding author states that there is no conflict of interest.

## Notes

### Competing Interest Statement

The authors have declared no competing interest.

### Author Declarations

(1) English Longitudinal Study of Ageing: our study used anonymised data from ELSA Wave 8, which is available to researchers via the UK Data Service via an institutional log in, and we adhered to the conditions of use. The ELSA study has ethical approval and participant consent as follows: 'This study involves human participants and the English Longitudinal Study of Ageing (ELSA) Wave 8 received ethical approval from the South Central - Berkshire Research Ethics Committee on 23 September 2015 (15/SC/0526)' (https://www.elsa-project.ac.uk/ethical-approval). Participants gave informed consent to participate in the study before taking part. (2) The study was approved by the Clinical Practice Research Datalink Independent Scientific Advisory Committee, protocol 23_002557. We adhered to the conditions of use and did not attempt to link between datasets.

### Summary of Updates

This version of the manuscript incorporates minor revisions in response to comments from reviewers.

