## Supplementary Materials for "Measuring Frailty: A Comparison of the Cumulative Deficit Model of Frailty in Survey and Routine Data"

**Table 1.** *Prevalence of Deficits in ELSA and CPRD, standardised by age and sex*

| eFI Item (CPRD) | CPRD (N) | CPRD (%) | ELSA Variable Description | ELSA (N) | ELSA (%) |
|---|---|---|---|---|---|
| **Deficits with similar definitions** | | | | | |
| Arthritis | 393,976 | 24.2% | Arthritis (hedibar) | 2536 | 48.9% |
| Atrial fibrillation | 181,824 | 11.2% | Abnormal heart rhythm (hedimar) | 673 | 13.0% |
| Cerebrovascular disease | 137,754 | 8.5% | Stroke (hedimst) | 369 | 7.1% |
| Diabetes | 293,810 | 18.1% | Diabetes or high blood sugar (hedimdi) | 760 | 14.7% |
| Falls | 118,752 | 7.3% | Whether fallen down since last interview (hefla) | 1517 | 29.3% |
| Fragility fracture | 137,300 | 8.4% | Whether has fractured hip (hefrac) | 46 | 0.9% |
| Hearing impairment | 380,415 | 23.4% | Self-reported hearing (while using hearing aid if appropriate) (hehear*) | 1307 | 25.2% |
| Heart failure | 72,672 | 4.5% | Congestive heart failure (hedimhf) | 60 | 1.2% |
| Hypertension | 835,123 | 51.4% | High blood pressure (hedimbp) | 2380 | 45.9% |
| Osteoporosis | 201,745 | 12.4% | Osteoporosis (hedibos) | 595 | 11.5% |
| Parkinsonism & tremor | 22,606 | 1.4% | Parkinson's (hedibpd) | 61 | 1.2% |
| Sleep disturbance | 135,783 | 8.4% | Whether felt their sleep was restless during the past week (pscedc) | 1815 | 35.2% |
| Social vulnerability | 107,327 | 6.6% | Difficulty managing money, eg paying bills, keeping track of expenses (headlmo) | 164 | 3.2% |
| Visual impairment | 579,784 | 35.7% | Self-reported eyesight (while using lenses if appropriate) (heeye*) | 222 | 4.3% |
| **Deficits corresponding to multiple deficits** | | | | | |
| Mobility & transfer problems | 98,861 | 6.1% | Difficulty walking 100m (hemobwa) | 836 | 16.1% |
| | | | Difficulty climbing several flights of stairs without resting (hemobcs) | 1937 | 37.3% |
| | | | Difficulty getting up from chair after sitting long periods (hemobch) | 1368 | 26.4% |
| | | | Difficulty climbing one flight of stairs without resting (hemobcl) | 894 | 17.2% |
| | | | Difficulty bathing (headlba) | 507 | 9.8% |
| | | | Timed walk: whether had pain whilst walking (mmpain) | 419 | 9.1% |
| | | | Difficulty getting in and out of bed (headlbe) | 293 | 5.6% |
| | | | Difficulty walking across a room (headlwa) | 200 | 3.9% |
| Activity limitation | 10,929 | 0.7% | Difficulty reaching or extending arms above shoulder level (hemobre) | 606 | 11.7% |
| | | | Difficulty stooping, kneeling or crouching (hemobst) | 2288 | 44.1% |

| | | | | | |
|---|---|---|---|---|---|
| | | | Difficulty lifting or carrying weights over 10 pounds (4.54kg) (hemobli) | 1328 | 25.6% |
| | | | Difficulty pulling or pushing large objects (hemobpu) | 1035 | 20.0% |
| | | | Difficulty picking up a 5p coin from a table (hemobpi) | 361 | 7.0% |
| Requirement for care | 26,790 | 1.6% | Difficulty doing work around the house or garden (headlhg) | 946 | 18.2% |
| | | | Difficulty dressing, including putting on shoes and socks (headldr) | 715 | 13.8% |
| | | | Difficulty shopping for groceries (headlsh) | 527 | 10.2% |
| | | | Difficulty preparing a hot meal (headlpr) | 269 | 5.2% |
| | | | Difficulty using the toilet including getting up or down(headlwc) | 199 | 3.8% |
| | | | Difficulty eating, such as cutting up food (headlea) | 129 | 2.5% |
| | | | Difficulty making telephone calls (headlph) | 128 | 2.5% |
| | | | Difficulty taking medications (headlme) | 120 | 2.3% |
| Memory & cognitive problems | 99,716 | 6.1% | Whether correct day of month given (cfdatd) | 902 | 17.7% |
| | | | Whether correct month given (cfdatm) | 158 | 3.1% |
| | | | Whether correct year given (cfdaty) | 162 | 3.2% |
| | | | Whether correct day given (cfday) | 139 | 2.7% |
| | | | Dementia (hedibde) | 80 | 1.5% |
| | | | Alzheimer's (hedibad) | 31 | 0.6% |
| Ischaemic heart disease | 441,930 | 27.2% | Angina (hediman) | 214 | 4.1% |
| | | | Heart attack (hedimmi) | 351 | 6.8% |
| Respiratory disease | 527,120 | 32.4% | Asthma (hedibas) | 602 | 11.6% |
| | | | Lung disease (hediblu) | 347 | 6.7% |

**Unique deficits**

| | | | |
|---|---|---|---|
| Anaemia / haematinic deficiency | 248,562 | 15.3% | --- |
| Chronic kidney disease | 294,341 | 18.1% | --- |
| Dizziness | 259,648 | 16.0% | --- |
| Dyspnoea | 263,729 | 16.2% | --- |
| Foot problems | 18,997 | 1.2% | --- |
| Heart valve disease | 28,389 | 1.7% | --- |
| Housebound | 37,953 | 2.3% | --- |
| Hypotension/syncope | 160,480 | 9.9% | --- |
| Peptic ulcer | 63,609 | 3.9% | --- |
| Peripheral vascular disease | 68,055 | 4.2% | --- |
| Skin ulcer | 111,345 | 6.8% | --- |

| | | | | | |
|---|---|---|---|---|---|
| Thyroid disease | 114,001 | 7.0% | --- | | |
| Polypharmacy | 751,984 | 46.3% | --- | | |
| Urinary incontinence | 120,755 | 7.4% | --- | | |
| Urinary system disease | 598,479 | 36.8% | --- | | |
| Weight loss & anorexia | 84,047 | 5.2% | --- | | |
| --- | | | Difficulty sitting 2 hrs (hemobsi) | 673 | 13.0% |
| --- | | | Cancer (hedibca) | 413 | 8.0% |
| --- | | | Psychiatric condition (hedibps) | 507 | 9.8% |
| --- | | | Whether felt depressed much of the time during the past week (psceda) | 619 | 12.0% |
| --- | | | Whether felt everything they did during the past week was an effort (pscedb) | 1044 | 20.2% |
| --- | | | Whether was happy much of the time during the past week (pscedd) | 426 | 8.3% |
| --- | | | Whether felt lonely much of the time during the past week (pscede) | 619 | 12.0% |
| --- | | | Whether enjoyed life much of the time during the past week (pscedf) | 394 | 7.6% |
| --- | | | Whether felt sad much of the time duing the past week (pscedg) | 997 | 19.3% |
| --- | | | Whether could not get going much of the time during the past week (pscedh) | 1039 | 20.1% |
| --- | | | Self-reported general health (hehelf*) | 1519 | 29.3% |
| --- | | | Whether had joint replacement (heji) | 241 | 4.6% |
| --- | | | Difficulty using map to figure out how to get around strange place (headlma) | 228 | 4.4% |

* Unless otherwise indicated, ELSA deficits were binary (0 = no deficit is reported; 1 = deficit is reported).

*Ordinal variables were dichotomised from the reported 5-point Likert scale (Excellent, Very good, Good, Fair, Poor), where Fair or Poor were treated as deficit present.

**Table 2**. *Age-Sex Adjusted Odds Ratios for Deficits with Similar Definitions in ELSA and CPRD*

| eFI Deficit (CPRD) | ELSA Frailty Index Deficit | Age-Sex Adjusted OR (95% CI) | p-value |
|---|---|---|---|
| Arthritis | Arthritis | 0.320 (0.303-0.338) | 0.000 |
| Atrial fibrillation | Abnormal heart rhythm | 0.775 (0.713-0.842) | 0.000 |
| Cerebrovascular disease | Stroke | 1.148 (1.031-1.278) | 0.012 |
| Diabetes | Diabetes or high blood sugar | 1.259 (1.165-1.360) | 0.000 |
| Falls | Whether fallen down since last interview | 0.155 (0.146-0.165) | 0.000 |
| Fragility fracture | Whether has fractured hip | 10.382 (7.760-13.889) | 0.000 |
| Hearing impairment | Self-reported hearing (while using hearing aid if appropriate) | 0.857 (0.803-0.914) | 0.000 |
| Heart failure | Congestive heart failure | 3.773 (2.923-4.871) | 0.000 |
| Hypertension | High blood pressure | 1.218 (1.152-1.288) | 0.000 |
| Osteoporosis | Osteoporosis | 1.103 (1.010-1.205) | 0.030 |
| Parkinsonism & tremor | Parkinson's | 1.137 (0.883-1.465) | 0.319 |
| Sleep disturbance | Whether felt their sleep was restless during the past week | 0.165 (0.156-0.175) | 0.000 |
| Social vulnerability | Difficulty managing money, eg paying bills, keeping track of expenses | 2.065 (1.764-2.418) | 0.000 |
| Visual impairment | Self-reported eyesight (while using lenses if appropriate) | 13.499 (11.784-15.463) | 0.000 |

**Table 3**. *Results of logistic regression model predicting frailty values by age, sex and datasource, with interaction terms, with age at 65 as the intercept. The reference categories for the variables are as follows: sex (male), age (1-year increase) and datasource (ELSA).*

| | Coef | Std err | t | P value | 95% CI |
|---|---|---|---|---|---|
| Intercept | 0.0712 | 0.002 | 28.713 | 0.000 | [0.0664, 0.0761] |
| Age | 0.0046 | 0.000 | 26.791 | 0.000 | [0.0042, 0.0049] |
| Sex | 0.0241 | 0.002 | 9.818 | 0.000 | [0.0193, 0.0289] |
| Datasource | 0.0029 | 0.002 | 1.162 | 0.245 | [-0.0020, 0.0077] |
| Age * Sex | 0.0002 | 0.000 | 12.448 | 0.000 | [0.0002, 0.0003] |
| Age * Datasource | 0.0009 | 0.000 | 5.303 | 0.000 | [0.0006, 0.0012] |
| Sex * Datasource | -0.0163 | 0.002 | -6.628 | 0.000 | [-0.0211, -0.0115] |

**Figure 1**. *Frailty Scores by Age Group and Data Source*.

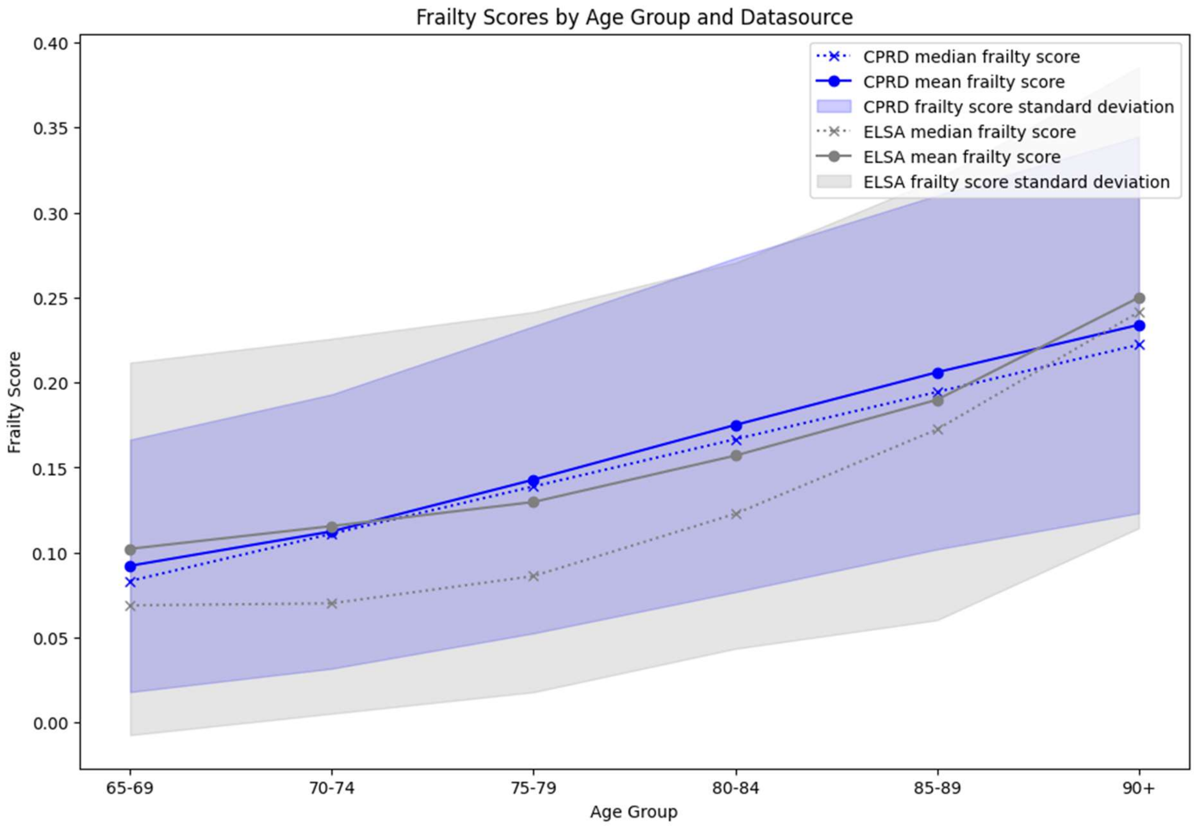


**Figure 2**. *Violin Plots of Frailty Index Scores by Sex, Age Group and Data Source*

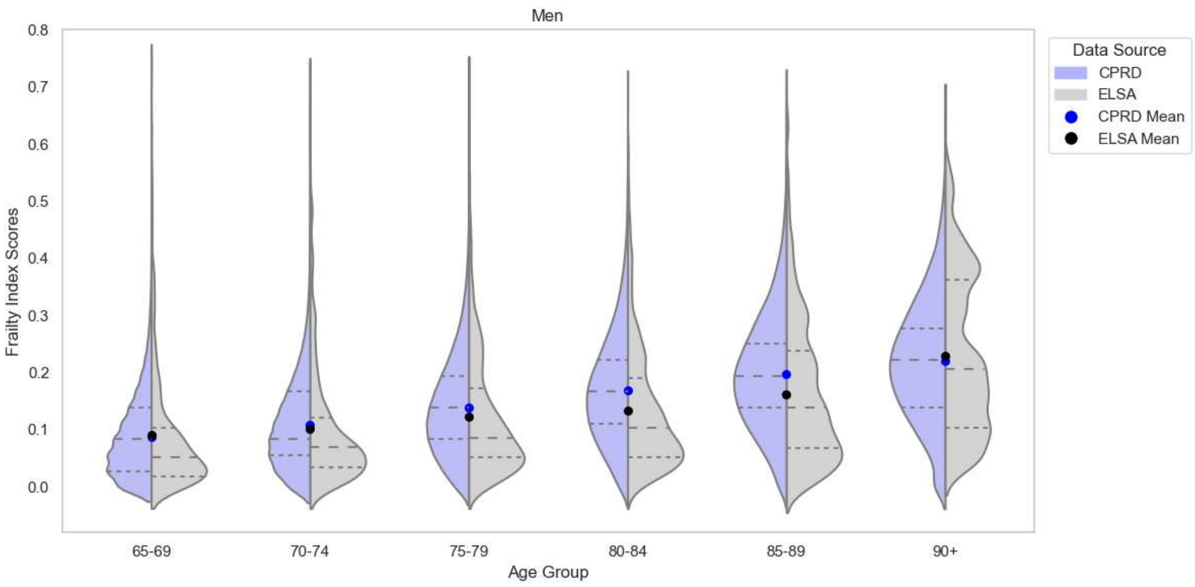

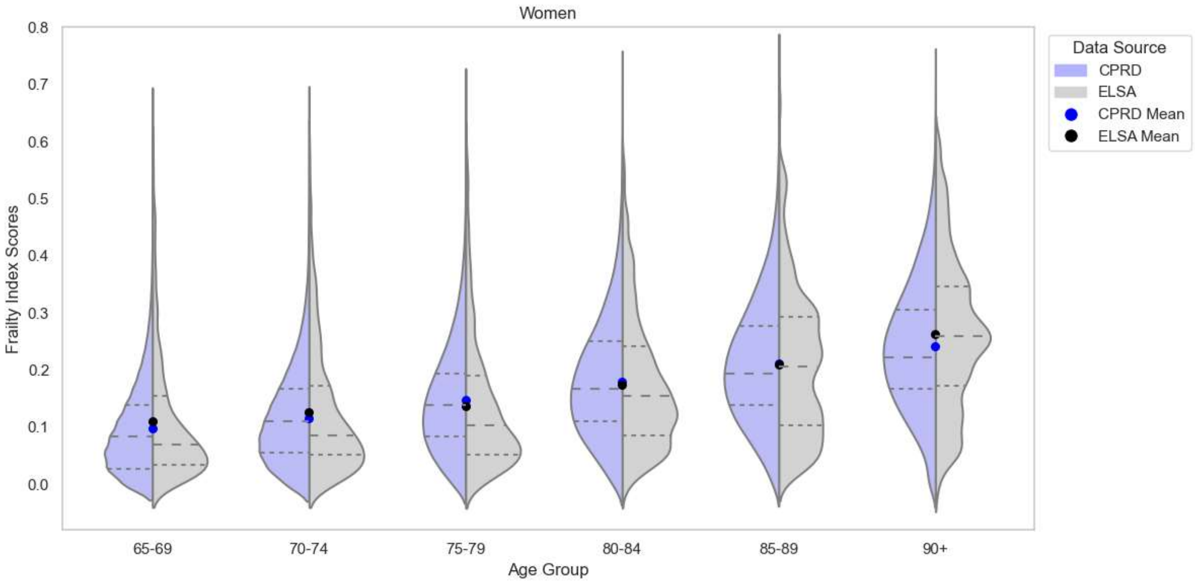


**Figure 3**. *Predicted frailty values by age, sex and datasource, with interaction terms*

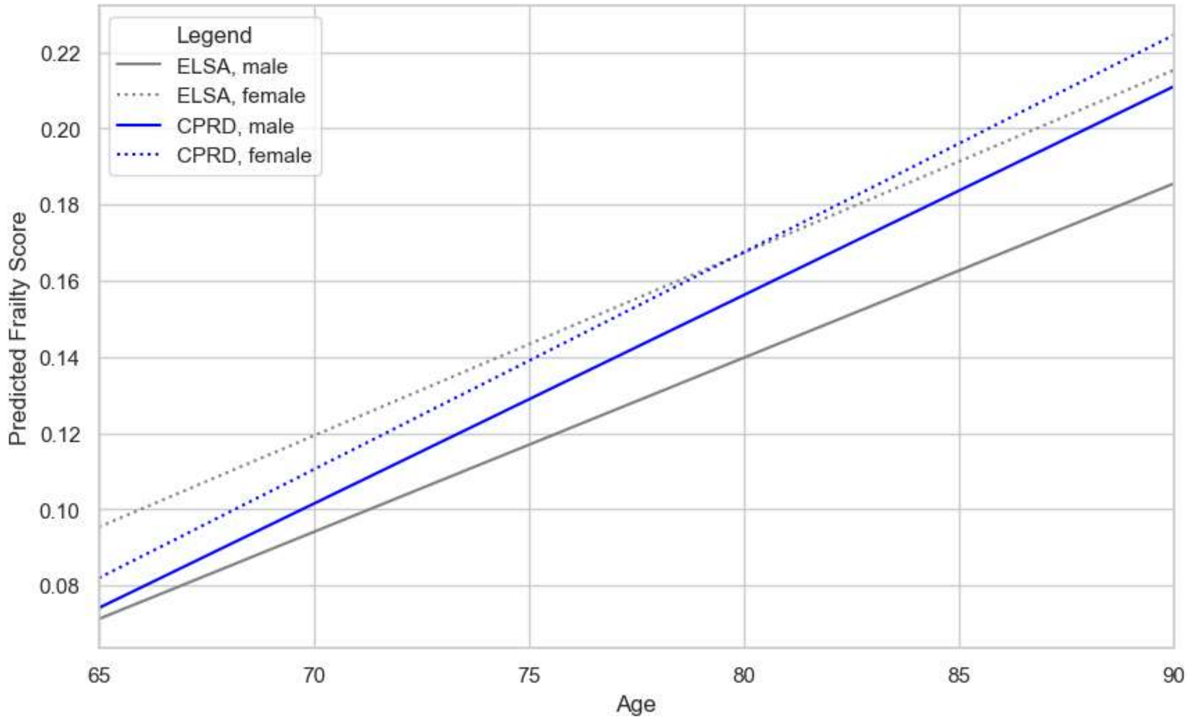
